# Bridging the Gap: Enhancing the Evaluation & Interpretation of Epidemic Forecasts for Researchers & Policymakers in Resource-Constrained Settings

**DOI:** 10.1101/2025.08.11.25333414

**Authors:** Paula Christen, Loice Achieng Ombajo, Anne Cori, Jeanette Dawa, Bimandra A Djaafara, Teresia Njoki Kimani, Camille MJ Schneider, Sabine L van Elsland, Mwangi Thumbi, Maria Veras, Charles Whittaker, Lilith K Whittles, Oliver J Watson

## Abstract

The COVID-19 pandemic highlighted the challenges of generating, interpreting, and acting on evidence in fast-moving, politically charged contexts. Epidemic forecasts became central to navigating uncertainty, yet their use, interpretation, and communication varied widely across settings. We conducted a global mixed-methods study, combining an online survey with 143 participants from 46 countries and 13 semi-structured interviews with individuals involved in COVID-19 policy dialogues. Forecasts informed a range of policy questions, from estimating epidemic size and health system needs to planning interventions, but their perceived value depended on clarity, contextual relevance, and timeliness. Decision-makers in high-income settings often used forecasts to explore scenarios and quantify uncertainty, supported by stronger modelling capacity, while counterparts in low- and middle-income countries emphasized the role of expert briefings and locally tailored insights in the face of limited baseline data and modelling infrastructure. Across contexts, forecasts were most actionable when they informed binary choices or compared concrete scenarios. Barriers to uptake included late delivery, lack of contextual fit, and limited technical familiarity, especially in resource-constrained settings. Strengthening forecast impact in such contexts will require modular, user-oriented tools, embedding modellers in response teams, co-developing decision-relevant metrics, and investing in foundational health data systems. These strategies can help ensure that forecasting is both technically robust and operationally relevant for future public health emergencies.

## INTRODUCTION

Evidence-informed policymaking is often described as a rational, staged process, where evidence is generated, rigorously appraised, synthesized, and used to inform decision-making. In practice, in fast-moving public health emergencies, this process is rarely linear or tidy. The COVID-19 pandemic highlighted that in crisis situations, evidence gathering, decision-making, and intervention often occur simultaneously. These overlapping processes are further complicated by compressed timelines, significant uncertainty, and highly charged political and social contexts.^1^

Early in the COVID-19 pandemic, decisions were often based on incomplete data^2^, past experiences (e.g., with severe acute respiratory syndrome or Middle East respiratory syndrome)^3,4^, expert opinion^5^, real-time international comparisons^6^, and epidemiological models.^7^ Epidemic forecasting, a subset of epidemiological modeling focused on short-term, probabilistic projections, emerged as a key tool for supporting decisions under uncertainty. Forecasts enabled rapid testing of policy hypotheses that would otherwise be costly, impractical, or unethical to explore.^8,9^ They were developed to inform decisions across sectors and levels, from hospital resource allocation to national strategies on pharmaceutical, and non-pharmaceutical interventions (NPIs) as well as international guidelines.

However, the availability and uptake of epidemiological forecasts in policy were uneven across countries and decision-making bodies.^10^ While efforts to build technical capacity in low- and middle-income countries are ongoing^11^, strengthening expertise alone will not close the science-policy gap. In many settings, models were developed without input from policymakers or a clear understanding of the decisions they were meant to inform.^12^ This disconnect between model development and the actual needs of policymakers was particularly evident during the COVID-19 pandemic and remains a central barrier to the effective use of forecasting in public health decision-making.

To support the timely and effective use of epidemic forecasts in public health emergencies— particularly in resource-constrained settings—this study examines the types of policy questions forecasts were used to address, the metrics and formats that shaped their utility, and the unmet needs that limited their impact. Drawing on experiences from the COVID-19 pandemic, it identifies recurring patterns that influenced uptake and interpretation, with the goal of informing how forecasts can be better aligned with decision-making needs in future crises. In doing so, this work complements ongoing efforts to promote data and statistical literacy among decision-makers^13^, while also calling attention to the role of the modelling community in shaping the policy relevance of their outputs.^12^

## METHODS

### Study Design

We employed a mixed-methods approach to examine the use and interpretation of epidemic forecasts in policy decision-making during the COVID-19 pandemic. Data were collected through an online survey and semi-structured follow-up interviews with stakeholders involved in the development, implementation, and evaluation of pandemic response policies and programmes.

### Survey

The online survey was developed using Qualtrics software and included 24 multiple-choice and open-ended questions. Survey items were informed by a review of the literature and input from one expert on modeling and policy engagement. The survey consisted of five sections covering participants’ engagement in COVID-19 policy dialogues, the types, and sources of epidemic forecasts accessed, policy questions addressed using forecasts, evaluation practices, and perceived credibility of forecasts, and barriers to their effective use in decision-making (Supplement 1A). The survey was translated into French and Spanish by native speakers and piloted with individuals experienced in modeling and policy engagement. Responses submitted in French or Spanish were translated into English for analysis.

Participants were eligible if they self-identified as having engaged directly or indirectly in COVID-19 policy dialogues. These dialogues were defined as decision-making processes related to the planning, development, or implementation of COVID-19 policies. Direct engagement referred to participation in meetings or discussions, while indirect engagement included the provision of evidence or technical advice to decision-makers.

Participants were recruited using convenience sampling through existing professional networks, snowball sampling, social media, and outreach during a satellite event at the Infectious Disease Modelling Conference, Bangkok, in November 2024. In addition, an email containing study information and a survey link was distributed using Microsoft Outlook Mail Merge software in December 2024. Upon clicking the link, participants were directed to a webpage containing a participant information sheet. Informed consent was obtained electronically before proceeding to the survey. The survey was open, meaning any eligible individual could participate. Participation was voluntary, and respondents had the option to remain anonymous, and to stop completing the survey at any point. The survey remained open for six months, from 23 September 2024 until 20 March 2025.

### Interviews

Semi-structured interviews were conducted via Microsoft Teams. Participants included a purposive sample of survey respondents who consented to follow-up, as well as additional individuals recruited specifically for the interview phase. Participants were selected to ensure diversity across regions, policy engagement types, and type of organization. Where preferred, interviews were conducted in the participant’s native language and later translated into English for analysis. All interviews were audio-recorded with consent, transcribed using Microsoft Teams’ transcription software, and pseudonymised prior to analysis.

Interviews probed participants’ experiences using epidemic forecasts during the COVID-19 pandemic, including the types of forecasts they relied on, their perceived value, and the challenges encountered in their use. Participants were asked to reflect on the accessibility and usefulness of forecast metrics, the key policy questions forecasts addressed or failed to address, and the ways in which forecasts were evaluated before informing decisions. Additional questions explored perceived barriers to effective forecast use and solicited recommendations for improving forecast development, communication, and integration into policy (Supplement 1B).

### Analysis

#### Survey Data

Survey responses were exported and analysed using R. Descriptive statistics (counts and percentages) were used to summarise quantitative responses. Analyses were stratified by stakeholder type and World Bank income classification, with a primary comparison between economic development contexts. Analyses used different denominators, corresponding to the number of participants who answered each question, to account for item non-response and potential dropouts during the survey.

All source code used for data analysis is available on GitHub (https://github.com/paulachristen/infectech_manuscript).^14^

#### Qualitative Data

Interview transcripts and responses to open-ended survey questions were analysed thematically using the Framework Method in NVivo (version 15). Coding was conducted deductively, examining differences by the level of policy engagement (e.g., field-level, subnational, national) and economic development context.

To ensure consistency and reduce bias, data were double-coded independently by two researchers (PC and SvL), and coding discrepancies were resolved through discussion. Once sufficient inter-coder agreement was achieved (kappa > 0·75 ^15^), the remaining interviews and open-text responses were coded using the shared codebook. Themes were refined iteratively through memo writing, constant comparison across interviews, and collaborative team discussions.

#### Ethics

This study was conducted in compliance with the <u>CHERRIES guidelines</u>. Ethical approval was obtained from the Imperial College Research Ethics Committee (ICREC reference number: 7095429). Participation was voluntary, and individuals could withdraw at any stage prior to data analysis. All data were pseudonymised and stored securely in accordance with institutional data protection policies.

## RESULTS

A total of 143 individuals enrolled in the online survey. Of these, 73·4% (105/143) reported involvement in COVID-19 policy dialogues at the national, sub-national, or field levels across 46 countries. Respondents reported working across multiple geographic levels: 19·6% (28/143) at the global level, 26·6% (38/143) at the regional level, 67·8% (97/143) at the national level, 36·4% (52/143) at the sub-national level, and 25·9% (37/143) at the field or community level. By country income classification, 13·3% (19/143) worked in low-income countries (LICs), 35·0% (50/143) in lower-middle-income countries (LMICs), 9·8% (14/143) in upper-middle-income countries (UMICs), and 23·1% (33/143) in high-income countries (HICs) during the COVID-19 pandemic. Further details on survey participation by section are provided in Supplement 2, Figure 1. In addition, thirteen participants took part in in-depth interviews, of which eleven were included in the analysis, having directly or indirectly engaged with epidemic forecasts (Supplement 2, Table 1).

**Figure 1.**
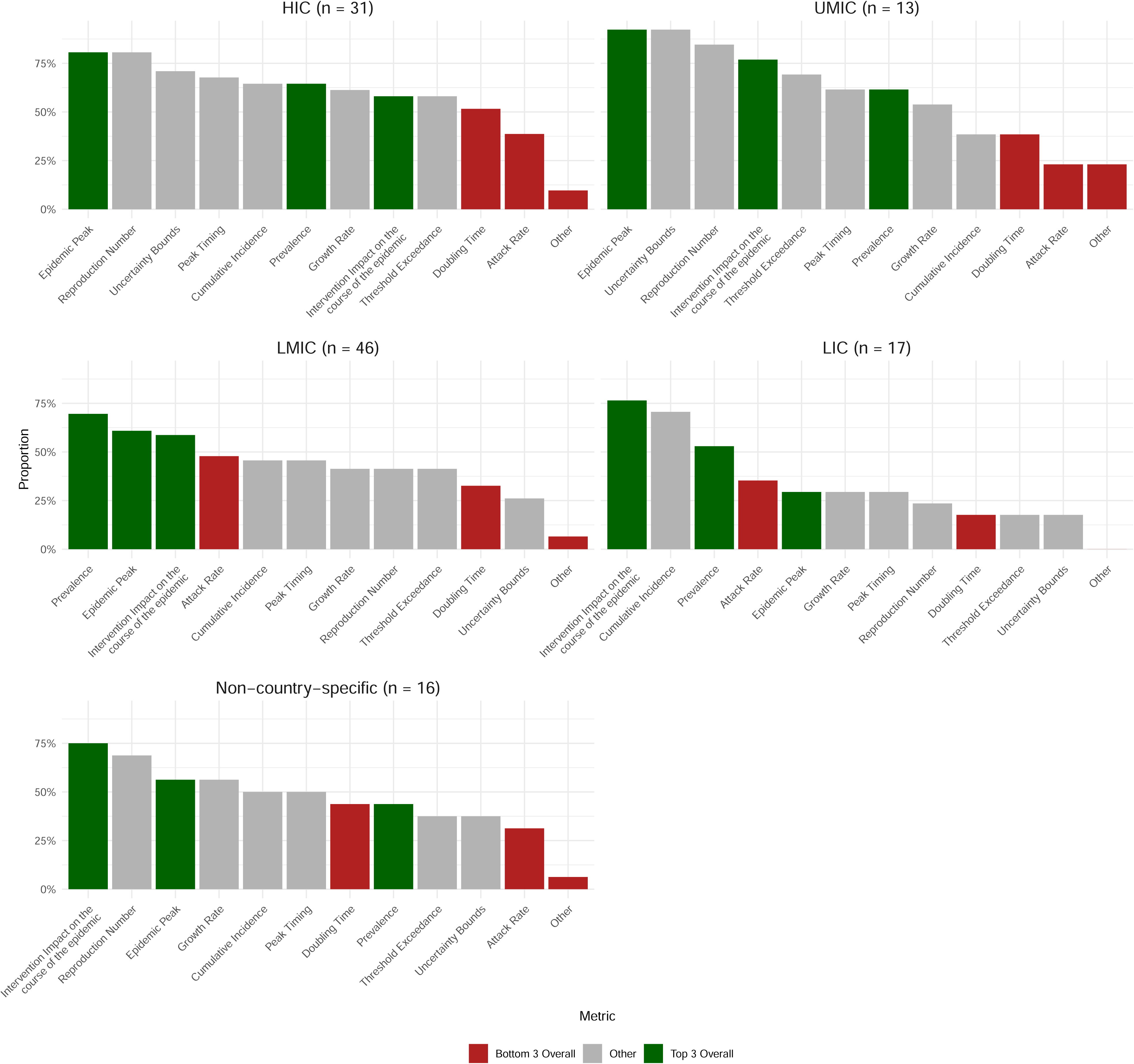
Forecasting metrics communicated during COVID-19 policy dialogues, by country income group. Data are drawn from a survey in which respondents were asked: “From the following list of numerical features, which ones did you communicate to your supervisor(s) or in policy dialogues with groups managing the COVID-19 pandemic response?” Responses are shown as percentages, stratified by country income group. Green bars represent the three most frequently reported metrics selected by all survey respondents, red bars the three least reported, and grey bars all others. Income group classifications follow the World Bank definitions: HIC = high-income countries; UMIC = upper-middle-income countries; LMIC = lower-middle-income countries; LIC = low-income countries.

**Table 1.**
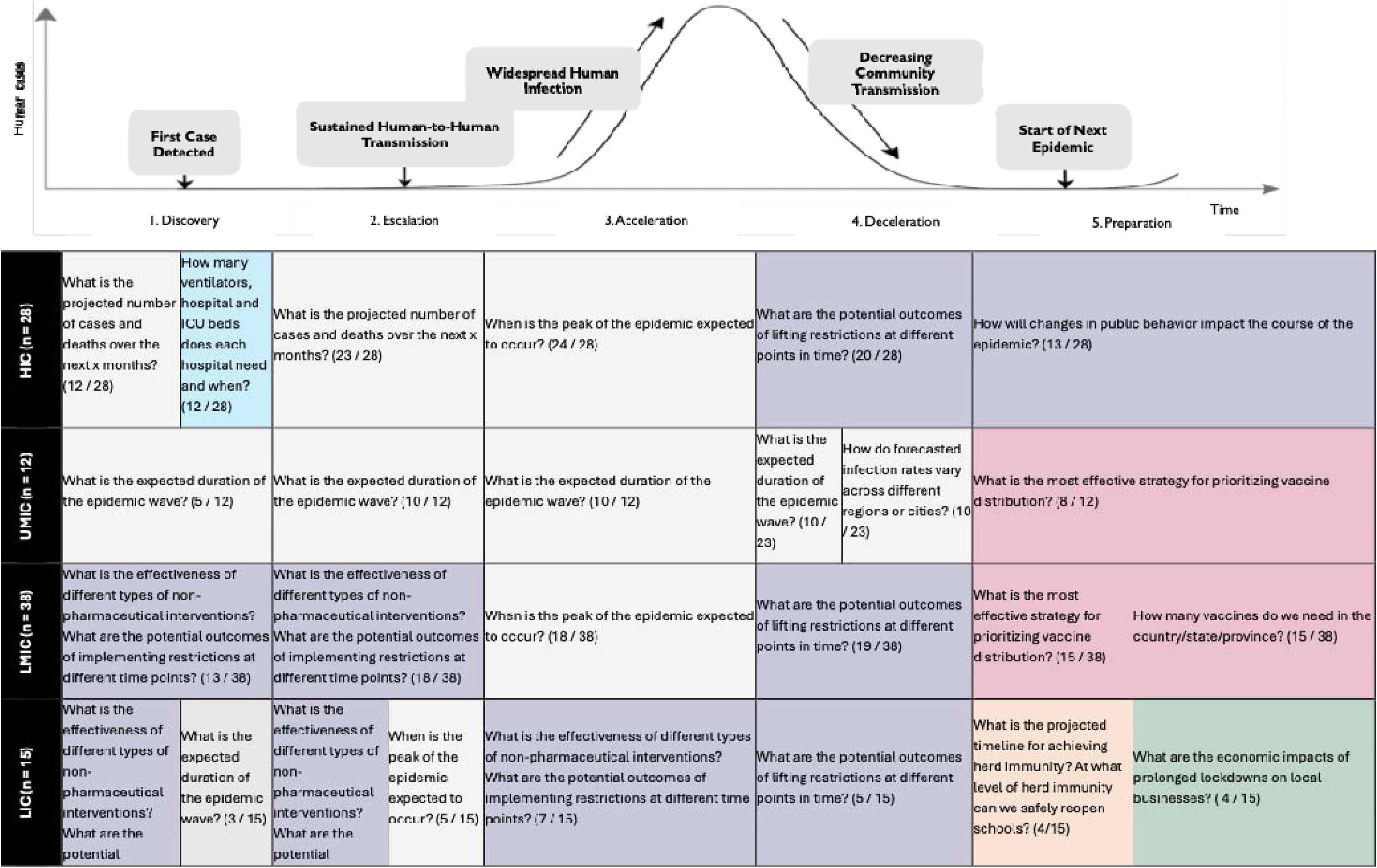
Most commonly selected forecast questions by pandemic phase and country income group. This table presents the most frequently selected responses to the survey question: “For each pandemic phase you were involved in, what questions did you or your supervisors use epidemic forecasts to help answer?” Responses are grouped by pandemic phase and country income category, classified using the World Bank categorization (2023): HIC = high-income country; UMIC = upper-middle-income country; LMIC = lower-middle-income country; LIC = low-income country. Other abbreviations: ICU = intensive care unit. Question categories are color-coded as follows: purple = Non-Pharmaceutical Interventions and Their Impact; grey = Epidemic Projections and Modeling; pink = Vaccination Strategies; orange = Herd Immunity and Reopening Strategies; blue = Healthcare Resource Allocation; green = Economic Impact.

### How Forecasts Were Used

#### Policy questions addressed by forecasts

Epidemic forecasts were used to inform six key types of policy questions: NPIs and their impact, herd immunity and re-opening strategies, economic impact, healthcare resource allocation, vaccination strategies, and epidemic projections and modelling (Table 1). These questions shifted over the course of the pandemic, reflecting evolving policy priorities. Early forecasts were used to estimate epidemic size and model the effectiveness of NPIs. As case numbers accelerated, decision-makers sought to anticipate peaks and healthcare demands. In later stages, attention turned to the implications of lifting restrictions and behavioural change.

> “Before the pandemic, the policy solution was generally to crush the epidemic - reduce infections and hospitalizations to zero. After the pandemic began, the objective shifted: policymakers needed to understand not just what the epidemic curve would look like without intervention, but how different policies could shape that curve, balancing health outcomes with costs, education, and broader societal impacts.” (ID110)

#### Variation by setting and level

At the global level, forecasts primarily informed high-level coordination and regional comparisons. A respondent from an international organization noted:

> “A lot of our role was reporting upwards… giving a global overview of trends. It was very high level… more like: ‘In this region of the world, cases are increasing very quickly,’ or ‘cases are stable,’ or ‘we don’t really know what’s going on.’” (ID102)

At national and subnational levels, forecasts were used to anticipate epidemic size and timing, assess health system readiness, and guide contingency planning. One respondent explained how they “projected the disease pattern, mortality, and morbidity… calculated daily positivity rates, and forecast where we might land in two or three months” (ID105). In HIC and LMIC, forecasts also supported immediate planning, enabling policymakers to make informed decisions on resource allocation and capacity management, including whether to scale up testing, procure vaccines, and drugs, or reallocate hospital infrastructure (ID103, ID111). In Iceland, a HIC with a small, geographically concentrated population, a senior public health official (ID113) emphasized how local conditions shaped their use of forecasts. Without in-house modelling capacity, they relied on a university team to model epidemic trajectories and hospital admissions. These projections—communicated visually through epidemic curves—helped justify decisions to rapidly expand ICU capacity and informed public health regulations. At the same time, the respondent noted limitations in interpreting international models in their setting, where small case numbers and high day-to-day variation made local data difficult to work with. “We are so few,” they explained, “our numbers are always like this—like a sawtooth.” (ID113)

In LIC, forecasts also supported geographically and economically targeted responses. In Malawi, they guided the location of treatment centres and the delivery of support to vulnerable groups such as vendors and truck drivers during lockdowns (ID104). Geographic granularity enabled decision-makers to identify “townships that would be more affected than others” (ID104) and allocate resources accordingly. In Madagascar, daily forecasts were used to adjust ambulance deployment and manage patient flows based on projected demand (ID109).

#### Decision-relevant metrics

In parallel with the evolution of policy questions, the specific metrics used to inform decisions shifted over time and across settings. Survey responses highlight that the most frequently communicated metrics included the projected impact of interventions (65·0%; 80/123), the epidemic peak (64·2%; 79/123), and the prevalence (62·0%; 76/123) (Figure 1).

However, usage patterns varied by setting. The size of the epidemic peak (e.g., the expected maximum number of daily hospital admissions) was among the top three metrics shared in all income settings except LICs, where the projected impact of interventions was most commonly communicated. The reproduction number was frequently cited in HICs, UMICs, and at the global and regional levels, but was rarely cited by respondents in LICs. Doubling time was among the least communicated metrics across most settings, with the exception of regional and global levels. Similarly, the attack rate ranked among the least used in LICs, but was more frequently reported in UMICs, where it was the fourth most commonly communicated metric. In Australia, focus initially centred on ICU occupancy, but shifted over time to more operational metrics like hospitalizations and workforce absenteeism, particularly during the Omicron wave (ID113).

### How Forecasts Were Communicated

#### Formats and framing of forecasts

Forecasts were received in a range of formats, with graphs (48·3%; 59/123), interactive dashboards (39·9%; 49/123), and scientific papers (38·5%; 47/123) among the most common (Figure 2). Across both survey and interview data, visual and concise outputs, particularly graphs, dashboards, and policy briefs, were consistently preferred over more technical formats such as point estimates or statistical intervals. These latter formats were often viewed as difficult to interpret or apply in real time.

**Figure 2.**
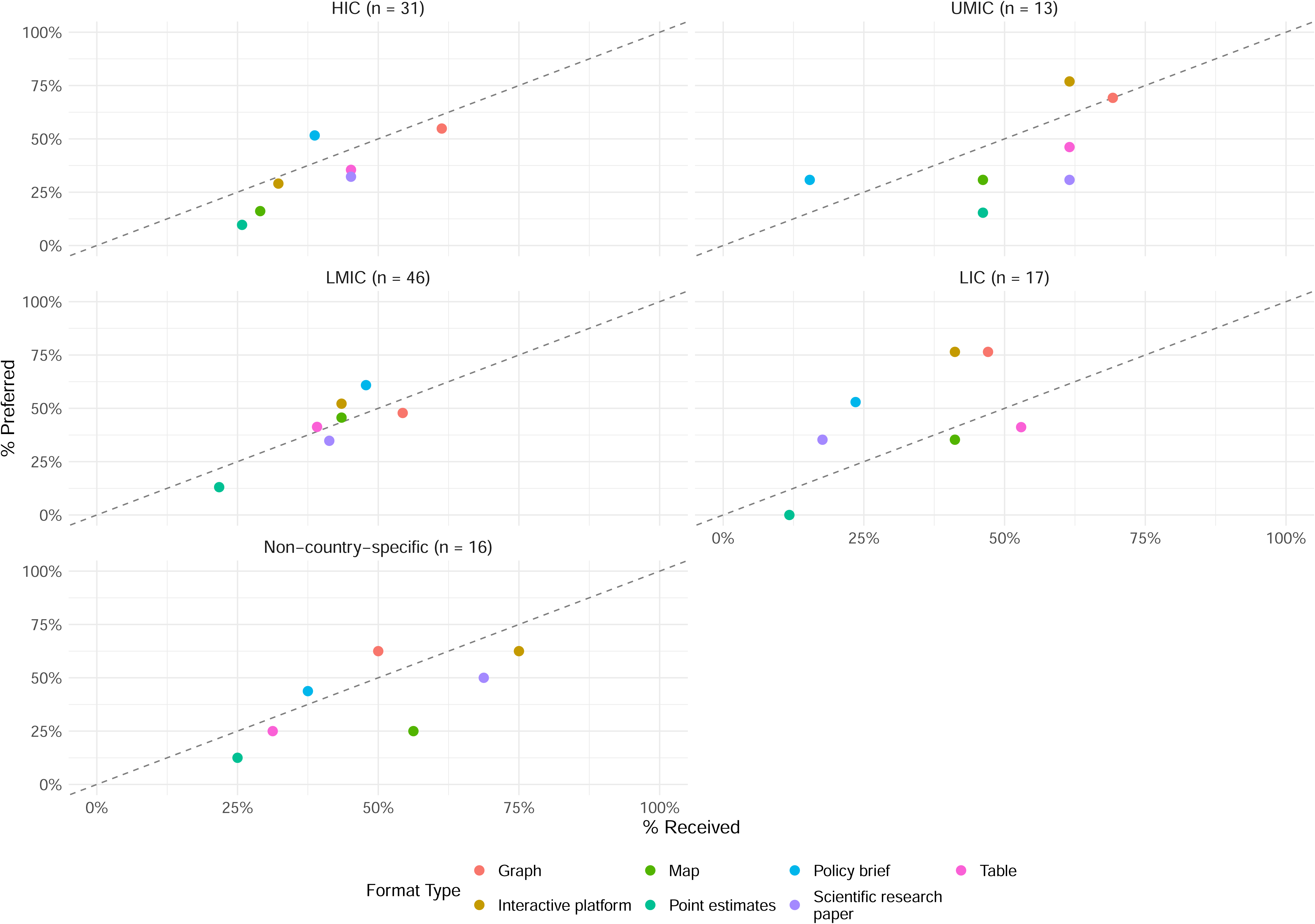
Alignment between received and preferred epidemic forecast formats, stratified by country income group. This figure presents responses to two survey questions: “In what format did you receive epidemic forecasts?” and “What is your preferred format to receive epidemic forecasts in?” Each point represents a format type, with its position showing the percentage of respondents who reported receiving (x-axis) versus preferring (y-axis) the format for epidemic forecasts. Points above the diagonal dashed line indicate formats that are under-provided relative to preference; points below the line indicate formats that are more often received than preferred. Panels are stratified by respondents’ country income group: HIC = high-income countries; UMIC = upper-middle-income countries; LMIC = lower-middle-income countries; LIC = low-income countries; and “Non-country-specific” for respondents without a clear national focus. Country income group classifications follow the World Bank definitions.

Forecasts were most impactful when used to assess whether key policy thresholds had been crossed, supporting binary decisions such as whether to scale up testing or approve vaccine orders. As one policymaker explained, “the issue is more about a binary decision, do I need to order more tests, do I need to sign off vaccines, rather than interpreting detailed point estimates” (ID110). This was echoed by another policymaker in Iceland (ID113).

Scenario-based presentations were highly valued across settings for conveying uncertainty and supporting adaptive decision-making. Respondents from Sweden (ID111), Zimbabwe (ID107), Lao PDR (ID103), and the UK (ID110) described using scenario comparisons to evaluate the implications of different actions, such as lockdowns, school closures, or changes in vaccine coverage.

#### Preferences for presentation and expert engagement

Interactive dashboards received mixed feedback. Some users appreciated their ability to explore scenarios and monitor trends, particularly in countries with limited in-country modelling capacity (ID101). However, their utility was dependent on users’ time, digital skills, and familiarity with the tools, factors that were cited as barriers in LIC settings such as Malawi (ID104).

Forecasts were most useful when accompanied by expert interpretation and interactive dialogue. Policymakers emphasized the value of real-time briefings that created space for questions and clarification, especially in LMICs and LICs where contextual understanding was crucial (ID110, ID107). In high-level policy settings, clarity, and simplicity were also key. One respondent explained that decision-makers often preferred narrative framings over technical details, asking: “Is it going to be twice as bad, half as bad, or about the same?” (ID113). While dashboards were valued by some subnational actors, they were rarely used in senior policy meetings; visual summaries and concise briefings were favored. Without engagement, technically sound forecasts were often seen as less helpful or even confusing (ID101).

While scientific papers were valued for their credibility, they were not considered well-suited to decision-making in real time. One respondent remarked, “[scientific papers] shouldn’t be the main tool… but the proof of work is sometimes very helpful” (ID110).

Among survey respondents in HIC and UMIC, 72·0% (18/25) and 83·3% (10/12), respectively, identified the explicit presentation of uncertainty as a key confidence factor, compared to just 34·3% (12/35) in LMIC and 23·1% (3/13) in LIC (Figure 3). Qualitative descriptors such as “high certainty” or “low certainty” were considered more intuitive according to a respondent at an international organization (ID102).

**Figure 3.**
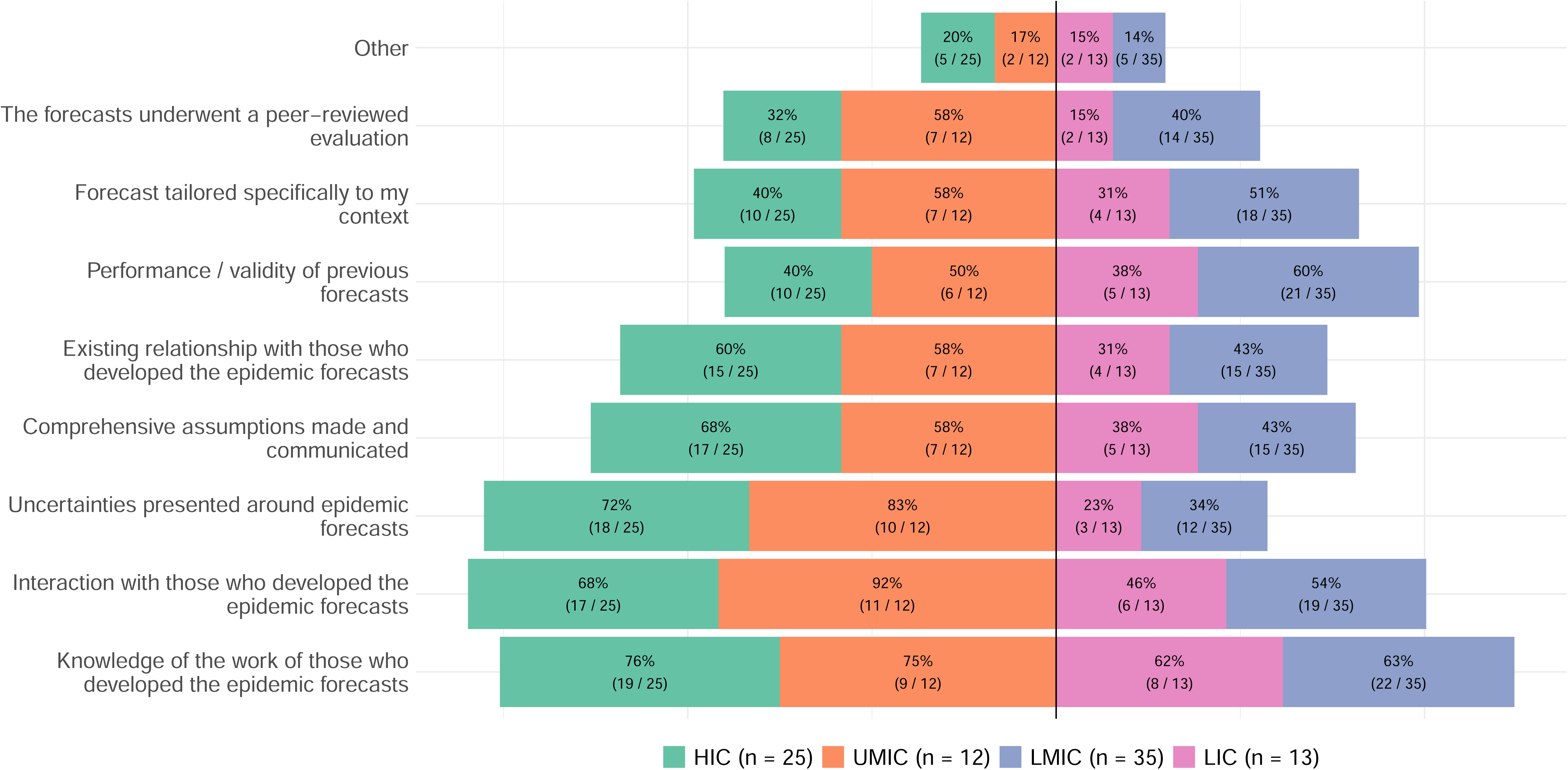
Factors contributing to confidence in using epidemic forecasts, by stakeholder group. This figure presents responses to the survey question: “What made you feel confident about informing your discussion or presentation to your supervisors with these epidemic forecasts?” Respondents selected one or more factors contributing to their confidence. The percentage of respondents selecting each factor is shown by the width of the bar, with the text showing also the number of respondents. Results are shown by country income group classified using the World Bank categorization (2023): HIC = high-income country (green); UMIC = upper-middle-income country (orange); LMIC = lower-middle-income country (violet); LIC = low-income country (pink).

### What Shaped Forecast Uptake

#### Barriers to integration

Barriers to the effective use of epidemic forecasts were most commonly rooted in foundational data limitations, issues of timing and delivery, challenges in interpretation, and misalignment with local contexts.

Unreliable, delayed, or incomplete data undermined the perceived credibility of forecasts; real-time decision-making was hindered when baseline data were missing or late. As one policymaker from Madagascar noted, “if the data had arrived on time, there wouldn’t have been forecasting problems” (ID109). Another added, “before the epidemic forecast, we need to have the baseline data” (ID105). Timing of forecast delivery also played a critical role. Forecasts that arrived too late, or without explanation, were less actionable. These issues were most frequently reported by respondents in UMICs (42%; 5/12) (Figure 4).

**Figure 4.**
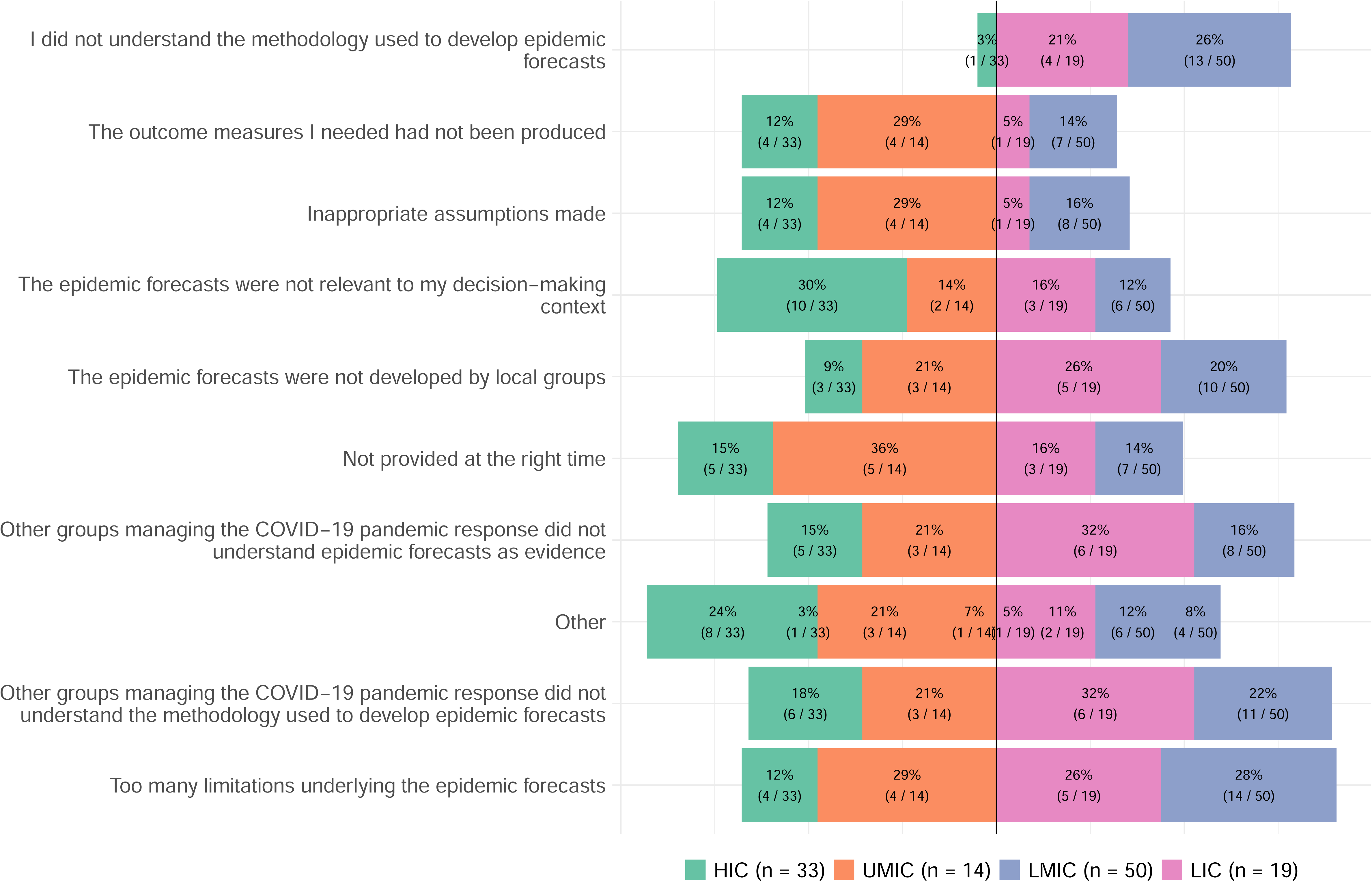
Barriers to using epidemic forecasts in policy dialogues during the COVID-19 pandemic, by country income group. Respondents were asked: “What hindered you from drawing on epidemic forecasts in dialogues with groups managing the COVID-19 pandemic response?” Results are shown by country income group classified using the World Bank categorization (2023): HIC = high-income country (green); UMIC = upper-middle-income country (orange); LMIC = lower-middle-income country (violet); LIC = low-income country (pink).

Several respondents noted that they or their colleagues did not fully understand the modelling approaches, which limited trust in and use of forecasts. Comprehension challenges among colleagues were particularly higher among LMIC and LIC respondents than HIC and UMIC respondents: 32% (11/34) of LMIC and 46% (6/13) of LIC participants reported that others involved in the response did not understand the modelling evidence. Self-reported gaps in understanding were also more common in these settings, 38% (13/34) in LMICs and 31% (4/13) in LICs, compared to just 4% (1/24) in HICs and 0% in UMICs (Figure 4).

A lack of locally developed models was another barrier. Nearly 30% of LMIC respondents (10/34) and 38% of LIC respondents (5/13) cited the absence of locally adapted tools or capacity as a reason forecasts were underused or mistrusted.

## DISCUSSION

To our knowledge, this is the first global, mixed-methods study to examine how epidemic forecasts were used, communicated, and perceived by public health decision-makers during the COVID-19 pandemic. Drawing on data from 46 countries across diverse economic contexts, the study addresses persistent gaps between the development of forecasts and their utility in decision-making in public health emergencies, particularly in resource-constrained settings. While forecasts were broadly recognised as valuable tools, their perceived utility depended on clear presentation, timely delivery, and alignment with operational needs.

Forecasts were most useful when tailored to the specific questions policymakers have. Although lists of potential policy questions have been proposed^16^, these have rarely been developed collaboratively with end-users.^17^ Our findings confirm that evidence needs are highly dynamic and context-specific, shaped by institutional roles, the stage of response, and existing decision-making structures.^13^ Survey responses provide a promising step toward structured anticipatory intelligence, reducing the need for modellers to “guess” what is useful for policymakers. A particularly novel finding was the heterogeneity in preferred forecast metrics across settings, reflecting differences in decision-making structures and technical capacity.

Tools presenting metrics relevant to users’ decision-making context and that speak to cognitive preferences, rather than offering one-size-fits-all formats, can enhance knowledge translation. Developing interactive, modular tools that accommodate different user types (e.g., national vs. subnational decision-makers) and support adaptive visualisation can make forecasts more policy-relevant. However, through our survey, we also found that digital skills and familiarity with the tools were some of the barriers to using forecasts during health emergencies. Hence, development and training of such tools need to be done as part of pandemic preparedness strategies to prepare the users on utilising the tools better during emergencies. Modellers can also draw on emerging evidence around forecast visualization—such as preferences for uncertainty display and layout clarity—to enhance the accessibility and policy relevance of their outputs.^18^

We found several barriers to the integration of forecasts to policymaking during health emergencies. New technologies may have potential to overcome these barriers for future usages. Emerging artificial intelligence (AI) methods offer promising tools to support knowledge translation. AI-powered summarisation can help convert complex model outputs into clear, policy-relevant insights. Advances in time-series foundation models and ensemble forecasting also improve the speed and robustness of predictions, even in data-sparse settings.^19,20^ These tools are especially valuable in low-resource contexts where forecasts are developed externally but used locally.^21^ Open-source packages with templates for uncertainty bands, scenario plots, and policy triggers could help bridge communication gaps between modellers and decision-makers. Without such tools, poorly conveyed uncertainty risks eroding trust and reducing forecast utility.^22^ While technical improvements are critical, they rely on strong foundations. With global health funding cuts, sustaining core data infrastructure and custodianship is essential. Reliable, timely data forms the backbone of forecasts^9^, yet in many settings, especially lower income settings, data systems remain tied to short-term projects and international aid, making them vulnerable to collapse when funding ends.^23^ Without stable investment, data collection, quality assurance, and institutional memory erode, undermining even the most sophisticated models.^24^ When full-scale modelling efforts are not feasible due to funding constraints, protecting essential data systems is vital for ensuring relevant evidence during future public health crises.

However, having robust data is only part of the solution. We found that confidence in forecasts often rested less on statistical sophistication and more on interpersonal familiarity, institutional credibility, and shared understanding, particularly in LICs and LMICs. Ensuring that data is translated into timely, decision-relevant insights will still require formal mechanisms, such as data-sharing agreements, memoranda of understanding, and embedding modellers within surveillance units or emergency response centres. These approaches can foster trusted collaboration. Furthermore, when modellers are closely engaged with operational surveillance teams, they become more attuned to data limitations and better positioned to shape what is collected and how it is used.^25^

This study has several limitations. First, it is subject to recall bias, as participants were asked to reflect on decisions and interactions that occurred during the COVID-19 pandemic. Future research could mitigate this by embedding similar assessments within real-time epidemic responses or simulation exercises, such as the Polaris Exercise conducted by WHO to test the Global Health Emergency Corps. Second, non-response bias is likely. Key workers, who experienced considerable psychological strain during the pandemic^26^, may have been less willing to revisit those experiences. Furthermore, while we focused on individuals involved in feeding evidence into political decision-making, our findings may not fully capture the perspectives or priorities of politicians themselves. Future research should endeavour to include political decision-makers directly. However, accessing their perspectives remains challenging due to the small number of individuals at higher levels of decision-making, their limited availability, and the protective gatekeeping that often surrounds them.^27^ Third, although the survey achieved broad geographic reach, attrition over its course (Supplement 3) may have weakened the representativeness of responses in later sections. Finally, the study relied on self-reported involvement in policy dialogue, which we were unable to independently verify; not all participants may have had the same level of engagement in decision-making during the pandemic.

In conclusion, bridging the gap between epidemic forecasting and public health decision-making requires a fundamental shift in approach. Forecasting should be reimagined not just as a technical modelling task, but as a core component of decision support infrastructure. Pandemic preparedness involves pre-defining the types of decisions forecasts are meant to support, which in turn can inform data collection, modelling design, and communication strategies well before crises emerge. Crucially, the development of forecasting tools and the training of end-users must take place during periods of stability, so that these tools can be readily understood, trusted, and deployed when they are needed most.

Emergency conditions demand a rethinking of how evidence is produced, disseminated, and interpreted. While the principles of effective knowledge translation – timeliness, accessibility, interpretability, and transparency – are well-established^28–30^, they must be retooled for contexts where decisions are time-sensitive, information is partial, and the policy process is nonlinear. Operationalising these principles remains an urgent task for researchers, funders, and international institutions alike, building on efforts by groups such as WHO and others, to ensure that modelling serves not just the advancement of science, but public health.

## Data Availability

The authors cannot make the underlying datasets publicly available for ethical and legal reasons, particularly due to the sensitive information included. The code for the statistical models is available from our GitHub repository.

https://github.com/paulachristen/infectech_manuscript

## Author contribution statement

CW, LKW, OJW, BAD, and PC conceptualized the research motivation. OJW and PC designed the study. AC, TNK, and CW refined the data collection tools. CMJS translated data into English. JD, OJW, MV, and PC identified and recruited study participants. OJW, SvL, and PC analyzed the data. PC and OJW conceptualized the manuscript and drafted the initial version. All authors reviewed the results and contributed to the final manuscript.

## Acknowledgements

The authors are deeply grateful to all study participants who generously contributed their time and insights.

## Data sharing agreement

The authors cannot make the underlying datasets publicly available for ethical and legal reasons, particularly due to the sensitive information included. The code for the statistical models is available from <u>our GitHub repository</u>.

## Declaration of generative AI and AI-assisted technologies in the writing process

During the preparation of this work the authors used ChatGPT-4o in order to improve text clarity. After using this tool, the authors reviewed and edited the content as needed and take full responsibility for the content of the publication.

