## Supplement for "Bridging the Gap: Enhancing the Evaluation & Interpretation of Epidemic Forecasts for Researchers & Policymakers in Resource-Constrained Settings"

**Table of Contents**

**Supplement 1** ........ 2

**Supplement 2** ........ 19

**Supplement 1**

A. **Survey**



${e://Field/head}

**Block: Introduction**

**Si vous préférez répondre à l'enquête en français, veuillez suivre ce lien: https://imperial.eu.qualtrics.com/jfe/form/SV_9RHZ9aWVjK549Qa**

**Si prefiere responder a la encuesta en español, siga este enlace: https://imperial.eu.qualtrics.com/jfe/form/SV_2gY6rmeDQqdrz70**

Dear Participant,

We invite you to share your valuable insights on epidemic forecasting during the COVID-19 pandemic in this brief 10-minute survey.

Our goal is to bridge the gap between epidemic forecasts and practical decision-making, especially in resource-constrained settings. Your experiences will help us:

1. Identify unmet needs related to epidemic forecasting.
2. Develop user-friendly tools to evaluate and interpret forecasts for policymakers.

This survey is divided into five sections:

- **Participant Information:** To confirm your eligibility for this research.
- **Metrics:** To understand what types of epidemic forecasts were most commonly used in public health decision-making.
- **Questions:** To identify the key policy questions and decisions that epidemic forecasts helped address.
- **Evaluation:** To learn how epidemic forecasts were evaluated (if at all).
- **Barriers:** To uncover the main challenges that hindered the use and usefulness of epidemic forecasts for policymakers.

This research is conducted by Imperial College London, UK and the University of Nairobi, Kenya.

To learn more about your participation, please review the Participant Information Sheet. Before you proceed with the survey, we kindly request your online consent to participate in this study, please see below clauses and the consent form.

1. I confirm that I have read and understood the participant information sheet version 1.2 dated 29/07/2024 for the study titled *Bridging the Gap: Enhancing the Evaluation & Interpretation of Epidemic Forecasts for Researchers & Policymakers in Resource-Constrained Settings* and have had the opportunity to ask questions which have been answered fully.
2. I understand that my participation is voluntary and I am free to withdraw at any time, without giving any reason and without my legal rights nor treatment / healthcare being affected.
3. I understand that sections of any of my medical notes may be looked at by responsible individuals from Imperial College London, the NHS Trust or from regulatory authorities where it is relevant to my taking part in this research.
4. I give consent for information collected about me to be used to support other research or in the development of a new test by an academic institution or commercial company in the future, including those outside of the United Kingdom (which Imperial has ensured will keep this information secure).
5. I understand that data collected from me are a gift donated to Imperial College and that I will not personally benefit financially if this research leads to an invention and/or the successful development of a new test, product or service.
6. I consent to take part in Bridging the Gap: Enhancing the Evaluation & Interpretation of Epidemic Forecasts for Researchers & Policymakers in Resource-Constrained Settings.
7. I give consent to being contacted about the possibility to take part in other research studies.

○ I confirm to all of the above.

Thank you,

Paula Christen, Loice Achieng, Jeanette Dawa, Thumbi Mwangi, Charlie Whittaker, Lilith Whittles, Njoki Kimani, Maria Veras, Oliver Watson

For further information, please do not hesitate to contact Dr Paula Christen or Dr Oliver J Watson, who are leading this research study.

**Instruction**

*Please keep the following in mind while answering the survey questions:*

Throughout the survey, we frequently refer to

- *supervisors*. When answering the survey questions, consider supervisors to be those to whom you presented your work and who posed questions during the pandemic that needed answering. For example, this can be your line manager, who requested information from you; an organization that commissioned your work; or a task force group in which you discussed evidence.
- *epidemic forecasts.* Broadly, these are predictions of the future course of an infectious disease outbreak with which key characteristics such as the number of cases, geographic spread, and required hospital capacity can be estimated. Such forecasts can be done under hypothetical scenarios to explore the potential impact of different interventions or changes in disease transmission.

You can return to this page throughout the survey to remind yourself of this definition or to amend your responses.

**B1: Engagement in COVID-19 Pandemic Response**

**Which *one* organization best represents your primary place of work during the COVID-19 pandemic?**

For the rest of this survey, please answer all questions based on your experiences and role within this organization.

- ○ Government agency

- Independent national public health agency
- University/Research institution
- Non-government organization
- International organization (e.g, World Health Organization, other United Nations agencies)
- Private organization
- Hospital / Clinic / Other healthcare provider
- Other. Please specify:

**During the COVID-19 pandemic, did you contribute to policy dialogues with groups responsible for managing the pandemic?**

Examples of groups responsible for managing the pandemic at different operational levels are presented in the diagram below.

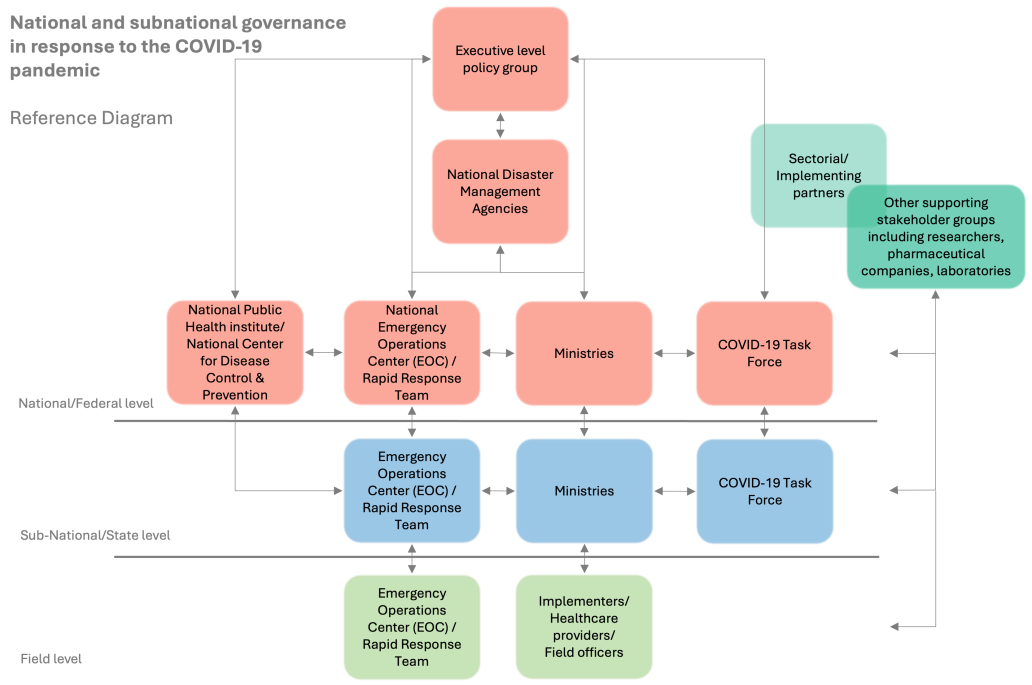

◯ Yes
◯ No

**Considering the reference diagram, which groups responsible for managing the pandemic did you engage with?**

Please select all that apply.

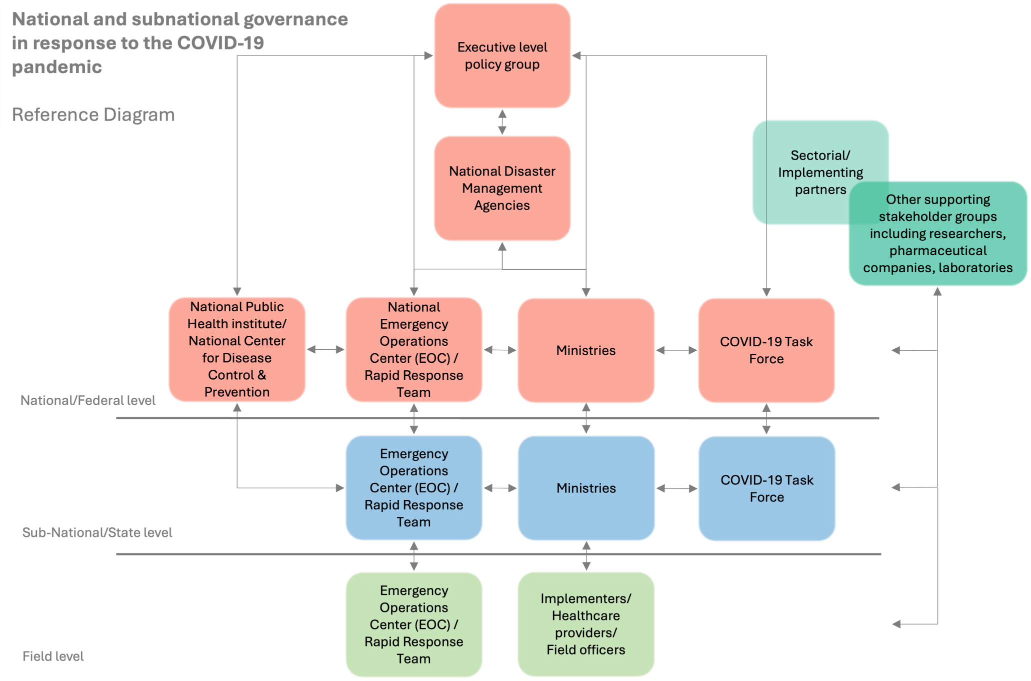


- ☐ Executive Level Policy Group
- ☐ National Disaster Management Agencies
- ☐ National / Federal Public Health Institute / National Centre for Disease Control & Prevention
- ☐ National / Federal Emergency Operations Center (EOC) / Rapid Response Team
- ☐ National / Federal Level Ministries
- ☐ National / Federal Level COVID-19 Task Force
- ☐ Sub-National / State Level Public Health Institute / National Centre for Disease Control & Prevention
- ☐ Sub-National / State Level Ministries
- ☐ Sub-National / State Level COVID-19 Task Force
- ☐ Field Emergency Operations Center (EOC) / Rapid Response Team
- ☐ Implementers / Healthcare providers / Field Officers

☐ Other supporting stakeholder groups including researchers, pharmaceutical companies, laboratories

☐ Sectorial / Implementing partners

☐ [ ] Other. Please specify:

Please note that the following question is similar to the previous question but focuses on the engagement of **your supervisor** in the COVID-19 pandemic response.

**During the COVID-19 pandemic, did you have supervisors, who contributed to policy dialogues with groups responsible for managing the pandemic?**

Examples of groups responsible for managing the pandemic at different operational levels are presented in the diagram below.

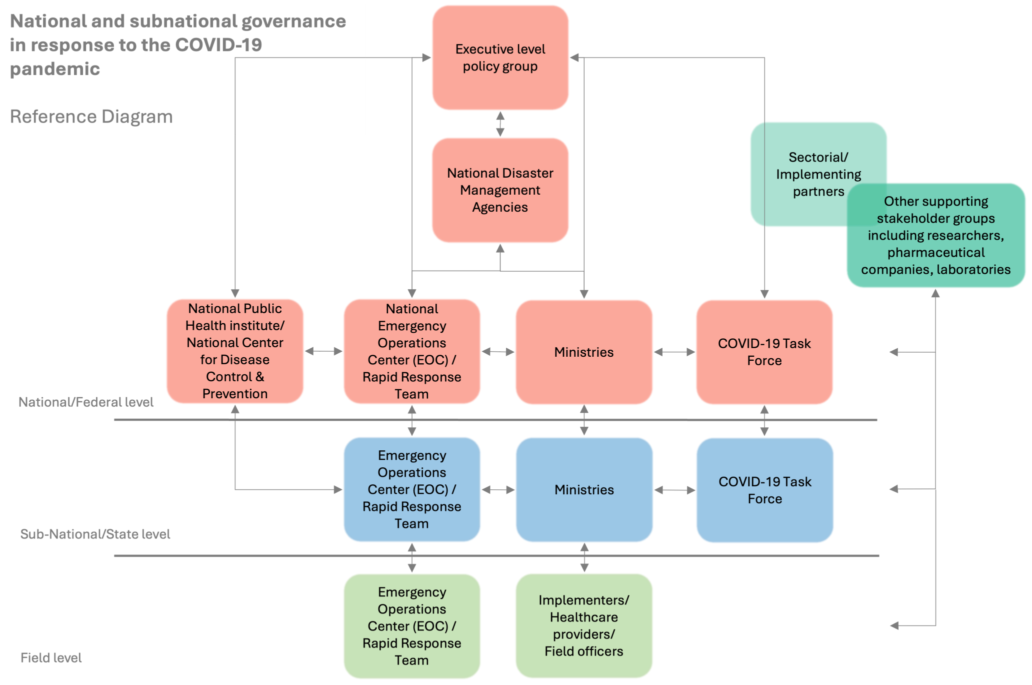


○ Yes

○ No

**Considering the reference diagram, which groups responsible for managing the pandemic did your supervisor(s) engage with?**

Please select all that apply.

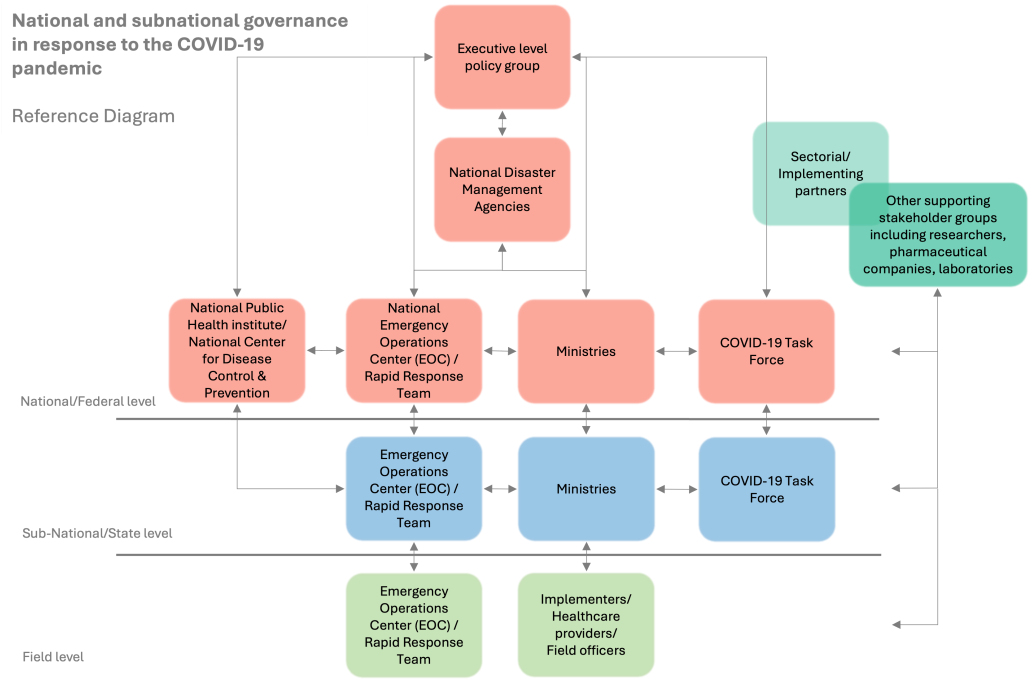


- ☐ Executive Level Policy Group
- ☐ National Disaster Management Agencies
- ☐ National / Federal Public Health Institute / National Centre for Disease Control & Prevention
- ☐ National / Federal Emergency Operations Center (EOC) / Rapid Response Team
- ☐ National / Federal Level Ministries
- ☐ National / Federal Level COVID-19 Task Force
- ☐ Sub-National / State Level Public Health Institute / National Centre for Disease Control & Prevention
- ☐ Sub-National / State Level Ministries
- ☐ Sub-National / State Level COVID-19 Task Force
- ☐ Field Emergency Operations Center (EOC) / Rapid Response Team
- ☐ Implementers / Healthcare providers / Field Officers
- ☐ Other supporting stakeholder groups including researchers, pharmaceutical companies, laboratories
- ☐ Sectorial / Implementing partners

☐ Other. Please specify: [ ]

**What best describes the geographic scope of your work during the COVID-19 pandemic?**

Please select all that apply.

- ☐ Global
- ☐ Regional (multiple countries)
- ☐ National
- ☐ Sub-national
- ☐ Local community

**Which country did most of your work focus on?**

[ ▼ ]

**Have you worked on responses to other health emergencies or disease outbreaks in low- or middle-income countries?**

- ◯ Yes
- ◯ No

***Optional*: Please provide your name and email address if you give consent for us to contact you for a follow-up interview.**

First Name [ ]

Last Name [ ]

Email Address [ ]

### B2: Epidemic Metrics

#### Did you receive or consult epidemic forecasts during the COVID-19 pandemic?

- ○ Yes
- ○ No

#### Where did you source epidemic forecasts from?

- ☐ Scientific literature (e.g. peer-reviewed publications, preprint platforms)
- ☐ Government and Health Agency Reports (e.g. WHO, COVID-19 Task Forces)
- ☐ Online Dashboards and Portals (e.g. IHME COVID-19 Projections)
- ☐ Press Releases and News Media (e.g. Online News Outlets)
- ☐ Social Media and Blogs (e.g. Twitter, Facebook)
- ☐ Technical Tools and Data Sharing Platforms (e.g. GitHub, Dropbox)

#### In what format did you receive epidemic forecasts?

- ☐ Map
- ☐ Table
- ☐ Graph
- ☐ Point estimates
- ☐ Interactive platform
- ☐ Policy brief (e.g., 2-page report with analysis)
- ☐ Scientific research paper
- ☐ Other. Please specify: ____________

#### What is your preferred format to receive epidemic forecasts in?

- ☐ Map
- ☐ Table
- ☐ Graph
- ☐ Single number / Point estimate
- ☐ Interactive platform
- ☐ Policy brief (e.g., 2-page report with analysis)
- ☐ Scientific research paper
- ☐ Other. Please specify: ____________

**Did you draw on any of these epidemic forecasts when presenting evidence to your supervisors or in policy dialogues with groups managing the COVID-19 pandemic response?**

- ○ Yes
- ○ No

**From the following list of numerical features, which ones did you communicate to your supervisor(s) or in policy dialogues with groups managing the COVID-19 pandemic response?**

- ☐ Epidemic Peak (e.g., the expected maximum number of daily hospital admissions)
- ☐ Threshold Exceedance (e.g., the date at which hospital occupancy is predicted to be greater than its capacity)
- ☐ Uncertainty Bounds (the range within which the true value is expected to lie, usually represented by confidence intervals or credible intervals)
- ☐ Peak Timing (e.g., the date when the peak number of cases is expected to occur)
- ☐ Cumulative Incidence (e.g., the probability that a person will develop a specific disease or condition over a certain period of time)
- ☐ Prevalence (e.g., the total number of active cases at a particular point in time)
- ☐ Reproduction Number (R0 or Rt) (the average number of secondary infections produced by an infected individual)
- ☐ Doubling Time (e.g., the time it takes for the number of cases to double)
- ☐ Growth Rate (e.g., the rate at which the number of cases is increasing or decreasing over time)
- ☐ Attack Rate (the proportion of the population that becomes infected in a time interval)
- ☐ Intervention Impact (the projected effect of specific interventions (e.g., social distancing, vaccination) on the course of the epidemic)
- ☐ [ ] Other. Please specify:

**B3: Questions**

**For each pandemic phase you were involved in (see diagram and phase definitions), what was the most pressing question you or your supervisors used epidemic forecasts to help answer?**

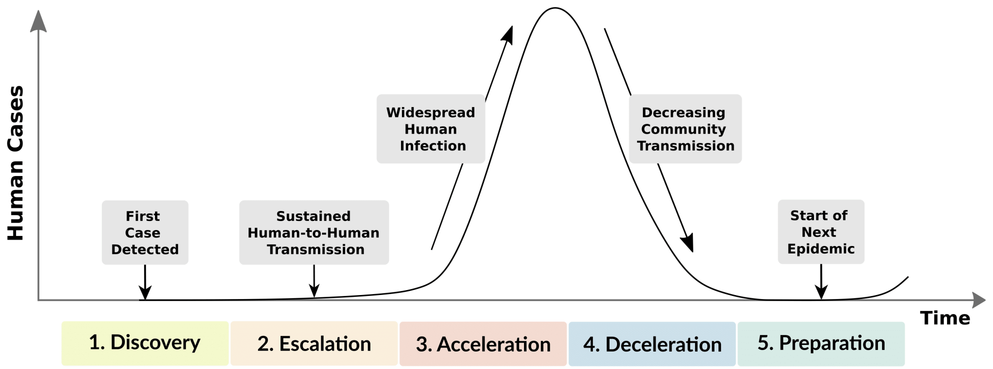


Please list below and indicate when, during the pandemic, these questions arose. Consider the following phases of the pandemic:

1. **Discovery:** The initial identification of the first human case.
2. **Escalation:** Increasing cases due to human-to-human transmission.
3. **Acceleration:** Significant increases in transmission due to widespread infection.
4. **Deceleration:** Decreasing cases after the peak in transmission.
5. **Preparation:** Limited transmission prior to possible next epidemic phase.

| | In each phase of the pandemic, what was the most pressing question you or your supervisors used epidemic forecasts to help answer? |
|---|---|
| Discovery | |
| Escalation | |
| Acceleration | |
| Deceleration | |
| Preparation | |

**For each pandemic phase you were involved in (see diagram and phase definitions), what questions did you or your superiors use epidemic forecasts to help answer?**

Please select the tick box if and when, during the pandemic, these questions arose. Consider the following phases of the pandemic in your country:

1. **Discovery:** The initial identification of the first human case.
2. **Escalation:** Increasing cases due to human-to-human transmission.
3. **Acceleration:** Significant increases in transmission due to widespread infection.
4. **Deceleration:** Decreasing cases after the peak in transmission.
5. **Preparation:** Limited transmission prior to possible next epidemic phase.

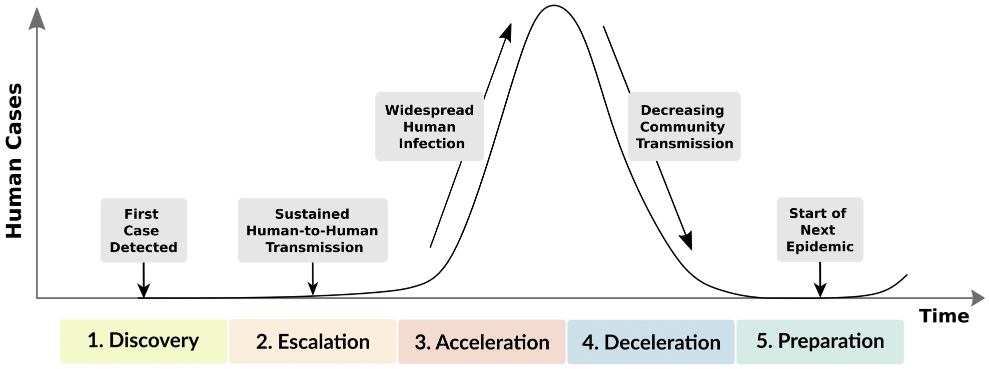


| | Discovery | Escalation | Acceleration | Deceleration | Preparation |
|---|---|---|---|---|---|
| What is the effectiveness of different types of non-pharmaceutical interventions? What are the potential outcomes of implementing restrictions at different time points? | ☐ | ☐ | ☐ | ☐ | ☐ |
| What are the potential outcomes of lifting restrictions at different points in time? | ☐ | ☐ | ☐ | ☐ | ☐ |
| What is the projected timeline for achieving herd immunity? At what level of herd immunity can we safely reopen schools? | ☐ | ☐ | ☐ | ☐ | ☐ |

| | Discovery | Escalation | Acceleration | Deceleration | Preparation |
|---|---|---|---|---|---|
| What is the projected number of cases and deaths over the next x months? | ☐ | ☐ | ☐ | ☐ | ☐ |
| When is the peak of the epidemic expected to occur? | ☐ | ☐ | ☐ | ☐ | ☐ |
| What is the expected duration of the epidemic wave? | ☐ | ☐ | ☐ | ☐ | ☐ |
| How do forecasted infection rates vary across different regions or cities? | ☐ | ☐ | ☐ | ☐ | ☐ |
| What are the economic impacts of prolonged lockdowns on local businesses? | ☐ | ☐ | ☐ | ☐ | ☐ |
| What is the most effective strategy for prioritizing vaccine distribution? | ☐ | ☐ | ☐ | ☐ | ☐ |
| How many vaccines do we need in the country/state/province? | ☐ | ☐ | ☐ | ☐ | ☐ |
| How many ventilators, hospital and ICU beds does each hospital need and when? | ☐ | ☐ | ☐ | ☐ | ☐ |
| How will changes in public behavior impact the course of the epidemic? | ☐ | ☐ | ☐ | ☐ | ☐ |

**Did you or your supervisors use epidemic forecasts to answer any other questions during the pandemic? If so, please elaborate on what these questions were and when they were posed.**

### B4: Evaluation

**Were you asked about the level of confidence or reliability in epidemic forecasts by your supervisors or in policy dialogues with groups responsible for managing the COVID-19 pandemic response?**

- ◯ Yes
- ◯ No

**How did you explain the level of confidence or reliability in epidemic forecasts to your supervisors or in policy dialogues with groups managing the COVID-19 pandemic response?**

**Were you asked about potential limitations or unknowns associated with epidemic forecasts to your supervisors or in policy dialogues with groups managing the COVID-19 pandemic response?**

- ◯ Yes
- ◯ No

**How did you present the potential limitations or unknowns associated with epidemic forecasts to your supervisors or in policy dialogues with groups managing the COVID-19 pandemic response?**

**What made you feel confident about informing your discussion or presentation to your supervisors with these epidemic forecasts?**

- ☐ Interaction with those who developed the epidemic forecasts
- ☐ The forecasts underwent a peer-reviewed evaluation
- ☐ Existing relationship with those who developed the epidemic forecasts
- ☐ Knowledge of the work of those who developed the epidemic forecasts
- ☐ Uncertainties presented around epidemic forecasts
- ☐ Comprehensive assumptions made and communicated
- ☐ Forecast tailored specifically to my context
- ☐ Performance / validity of previous forecasts

- [ ] Other. Please specify: ____

### B5: Barriers

#### What hindered you from drawing on epidemic forecasts in dialogues with groups managing the COVID-19 pandemic response?

- [ ] I did not understand epidemic forecasts as evidence
- [ ] Other groups managing the COVID-19 pandemic response did not understand epidemic forecasts as evidence
- [ ] I did not understand the methodology used to develop epidemic forecasts
- [ ] Other groups managing the COVID-19 pandemic response did not understand the methodology used to develop epidemic forecasts
- [ ] The epidemic forecasts were not relevant to my decision-making context
- [ ] The outcome measures I needed had not been produced
- [ ] Not provided at the right time
- [ ] The epidemic forecasts were not developed by local groups
- [ ] Inappropriate assumptions made
- [ ] Too many limitations underlying the epidemic forecasts
- [ ] Other. Please specify: ____

Accessibility

Powered by Qualtrics

### B. Interview guide

**Introduction**

a. Interviewer introduces themselves and their engagement in this work.
b. Explain background of research.
c. Aims of research:

Our goal is to bridge the gap between scientific forecasts and practical decision-making, especially in resource-constrained settings.

Your experiences will help us:

- Identify unmet needs related to epidemic forecasting.
- Develop user-friendly tools to evaluate and interpret forecasts for policymakers.

a. Explain what to expect in the interview.
b. Shortly reiterate information provided in survey.

---

**Interview**

**Section 1 – Reflections on the role of epidemic forecasts during the COVID-19 pandemic response**

i. How do you describe the role of epidemic forecasts during the COVID-19 pandemic response?
ii. Which types of epidemic forecasts (e.g., short-term case projections, hospital bed occupancy predictions, long-term scenario modeling) proved most valuable for decision-making? Why?
iii. What were the biggest challenges in using epidemic forecasts effectively during the rapidly evolving COVID-19 pandemic? Were there issues with data quality, model accuracy, or communication of results?
iv. How can we improve the development, communication, and utilization of epidemic forecasts to better prepare for future outbreaks?

**Section 2 – Reiteration of findings from survey**

So far, we have received xxx survey responses. Based on a preliminary analysis, we now better understand relevant metrics, questions, methods and requirements for evaluations of epidemic forecasts as well as barriers to using epidemic forecasts.

In the next section, we would like to probe these findings to understand whether these preliminary findings resonate with you and speak to your needs.

i. **Metrics: To understand the most accessible format of epidemic forecasts in public health decision-making.**
Confirmation:
  - Our survey suggests that [xxx] was the most accessible and useful format in which epidemic forecasts were presented/made available. Does this align with your experience? Why? Why not?
  - Our survey suggests that [xxx] were commonly communicated to supervisors or in meetings. Does this align with your experience? Why? Why not?

Value: Were these the most valuable types of forecast metrics for you, or were there others you found more useful? Why?

ii. **Questions: To identify the key policy questions and decisions that epidemic forecasts helped address.**
Relevance: Our survey identified the following as the top questions addressed by forecasts. [show and read printed list of key policy questions]
Did these questions align with your primary concerns during the pandemic?
Unmet Needs: Were there any critical policy questions that epidemic forecasts didn't address, but that you would have found valuable?
Priority: If you could only have forecasts for a limited number of questions, which would you prioritize and why?

iii. **Evaluation: To learn how epidemic forecasts were evaluated (if at all) before being used in decision-making.**

Practices: Our survey found that [summarize evaluation methods] were the most common ways forecasts were evaluated. Does this reflect your approach?
Satisfaction: Were they sufficient to assess forecast accuracy and reliability?
Ideal Evaluation: In an ideal scenario, how would you like epidemic forecasts to be evaluated before being used in decision-making?

iv. **Barriers: To uncover the main challenges that hindered the use and usefulness of epidemic forecasts for policymakers.**
Top Barriers: Our survey highlighted [list top barriers] as the main challenges to using forecasts effectively. Do these resonate with your experience?
Solutions: What steps could be taken to overcome these barriers and make forecasts more useful for policymakers like yourself?

**Wrap-Up:** Thank the participant for their time and valuable feedback.

**Supplement 2**

**Figure 1. Survey dropout rates by section and overall sample group.** The figure shows the percentage of respondents who dropped out of the survey at each section: Metrics, Questions, Evaluation, Confidence, and Barriers. Dropout rates are presented respondents' country income category, classified using the World Bank categorization (2023): HIC = high-income country; UMIC = upper-middle-income country; LMIC = lower-middle-income country; LIC = low-income country.

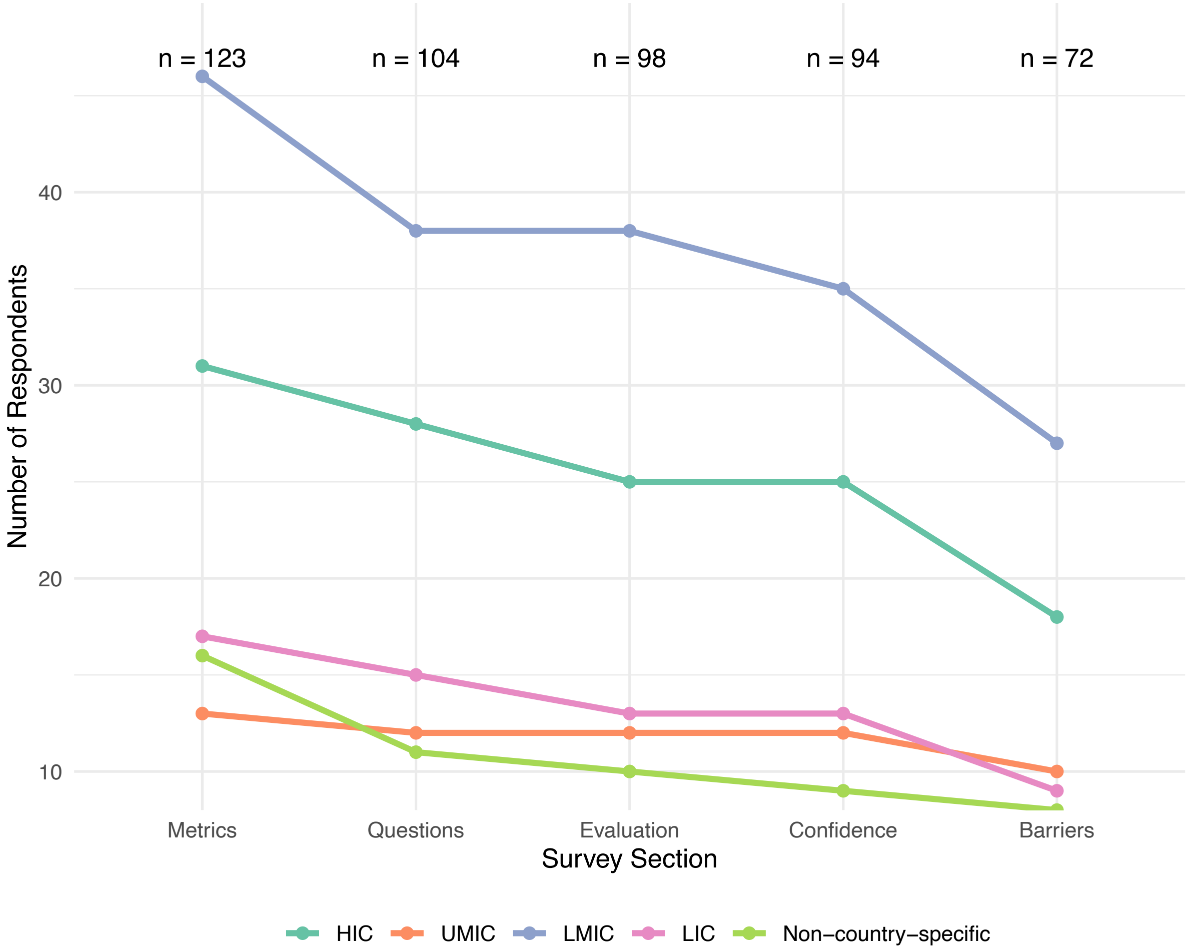

**Table 1 Characteristics of key informants participating in qualitative interviews.** The table summarizes the primary geographic scope of work, country income classification (LIC = low-income country, LMIC = lower-middle-income country, HIC = high-income country), organization type (NGO = non-governmental organization), and professional profile of 13 interviewees.

| ID | Primary geographic scope of work | Country income group | Type of organization | Profile |
|---|---|---|---|---|
| 101 | Regional (Europe) | | International organization | A senior infectious disease modeller working at a regional public health agency with a Europe-wide mandate during the COVID-19 pandemic. The participant led epidemic modelling efforts, coordinated cross-country forecast use, and supported high-level decision-making across multiple European countries. |
| 102 | Global | | International organization | An infectious disease modeller based at a multilateral public health agency with a global mandate. During the COVID-19 pandemic, they contributed to both high-level situational assessments and operational forecasting support for individual countries, primarily in the African region. Their work focused on producing in-house analyses using official data, and they emphasized the importance of timely, context-sensitive modelling outputs and clear communication of uncertainty to support decision-making. |
| 103 | Lao PDR | LMIC | Government agency | A senior public health official with decades of experience in infectious disease control and emergency preparedness. They have held leadership roles within a national health ministry and were closely involved in managing responses to major outbreaks, including SARS, MERS, and avian influenza. During the early stages of the COVID-19 pandemic, they coordinated national efforts and collaborated with international partners to develop scenario-based forecasts. These forecasts played a key role in informing preparedness planning and high-level policy decisions such as resource distribution and border control strategies. |
| 104 | Malawi | LIC | Government agency | A senior official at the Ministry of Health in Malawi, responsible for digital health and data quality. With a background in medicine and health systems strengthening, they were involved in the national COVID-19 response and worked closely with modelling teams to inform planning and resource allocation. |
| 105 | Pakistan | LMIC | Government agency | A senior government epidemiologist in Pakistan with leadership responsibilities for border health services and points of entry. With extensive training in field epidemiology and global health security, they played a key role in coordinating COVID-19 screening, response, and risk assessment. |
| 106 | Regional (Americas) | | International organization | A regional public health official working in Latin America and the Caribbean with responsibilities in emergency preparedness and outbreak response. During the COVID-19 pandemic, they used epidemic forecasts to inform supply planning and assess the potential impact of different public health strategies. |
| 107 | Zimbabwe | LMIC | Hospital / Clinic / Other healthcare provider | A public health leader with experience in national-level infectious disease modelling and policy advising. They co-led efforts to coordinate modeling inputs during the COVID-19 pandemic and collaborated with international partners to produce scenario-based forecasts. These forecasts informed government decision-making on public health measures such as lockdowns, school closures, and vaccination strategies. |
| 108 | Kenya | LMIC | NGO | A social development practitioner and policy advisor based in Kenya, involved in coordinating community-level COVID-19 response activities, including risk communication, service linkages, and support for gender-based violence survivors. |
| 109 | Madagascar | LMIC | Government agency | A regional public health official in Madagascar working in the environmental health department during the COVID-19 pandemic. Their responsibilities included overseeing hygiene, sanitation, and patient logistics across health facilities in the capital. They used short-term forecasts from a national health operations center to inform daily decisions on patient placement, ambulance routing, and resource allocation. |
| 110 | UK | HIC | Government agency | A senior modeller and public health advisor with responsibilities spanning infectious disease modelling, analytics, and policy engagement during the COVID-19 pandemic. They coordinated national-level forecasts and collaborated with academic institutions to inform real-time decisions on interventions, healthcare capacity, and broader societal trade-offs. |
| 111 | Sweden | HIC | Government agency | A senior public health official with a background in infectious disease epidemiology and leadership experience in national outbreak response. During the COVID-19 pandemic, they oversaw the development and communication of national epidemic forecasts, focusing on short-term hospital and ICU demand. Forecasts were presented to both government and healthcare system stakeholders using scenario- |

| | | | | |
|---|---|---|---|---|
| | | | | based graphs to communicate uncertainty. |
| 112 | Australia | HIC | University | An academic expert in epidemiology who led a national infectious disease modelling consortium during the COVID-19 pandemic. They contributed modelling analyses to high-level policy advisory groups, supporting critical public health decisions such as setting vaccine coverage targets for reopening, designing outbreak response strategies, and informing workforce planning during periods of high transmission. |
| 113 | Iceland | HIC | Government agency | The interviewee holds a senior position within a national public health agency in a high-income, small-population country. With a background in medicine, surgery, and biostatistics, she transitioned into public health leadership shortly before the COVID-19 pandemic. During the pandemic, she was centrally involved in national-level coordination, policymaking, and the implementation of emergency health measures, including border controls, digital contact tracing, and public guidance. |
